# Parsing Clinical Trial Eligibility Criteria for Cohort Query by a Multi-Input Multi-Output Sequence Labeling Model

**DOI:** 10.1101/2021.11.18.21266533

**Authors:** Shubo Tian, Pengfei Yin, Hansi Zhang, Arslan Erdengasileng, Jiang Bian, Zhe He

**Affiliations:** Department of Statistics, Florida State University; Department of Health Outcomes and Biomedical Informatics, University of Florida; School of Information, Florida State University

## Abstract

To enable electronic screening of eligible patients for clinical trials, free-text clinical trial eligibility criteria should be translated to a computable format. Natural language processing (NLP) techniques have the potential to automate this process. In this study, we explored a supervised multi-input multi-output (MIMO) sequence labelling model to parse eligibility criteria into combinations of fact and condition tuples. Our experiments on a small manually annotated training dataset showed that that the performance of the MIMO framework with a BERT-based encoder using all the input sequences achieved an overall lenient-level AUROC of 0.61. Although the performance is suboptimal, representing eligibility criteria into logical and semantically clear tuples can potentially make subsequent translation of these tuples into database queries more reliable.

## 1 Introduction

Randomized controlled trials are the gold-standard for evaluating the efficacy and safety of a treatment or intervention. Nevertheless, clinical trials often suffer from delayed patient accrual or insufficient participants, which may lead to early termination and cause significant financial loss for the sponsor. With the wide adoption of electronic health records (EHR), real-world EHR data allow us to evaluate the recruitment feasibility (Doods, Botteri, Dugas, & Fritz, 2014), perform electronic screening (Thadani, Weng, Bigger, Ennever, & Wajngurt, 2009), and assess the generalizability of the trials before enrollment (He et al., 2020). A necessary step to automate these analyses is to identify patients in the EHR data who satisfy the eligibility criteria of the trial, which are free-text sentences expressed in natural language and often with semantic ambiguities. It is thus important to extract key elements from eligibility criteria and translate them into computable database queries. Natural language processing (NLP) is a key technology to facilitate such translation.

Typically, parsing eligibility criteria consists of 5 major tasks: (1) sentence chunking, (2) named-entity recognition (NER) and concept mapping, (3) relationship extraction, (4) temporal constraint detection, and (5) negation detection. Depending on the specific techniques, some tasks (e.g., NER and relation extraction) can be done in a single joint model. Manual annotation of eligibility criteria is required for building a robust criteria parser but it is expensive, labor intensive, and requires clinical domain knowledge. Therefore, an open question is “*how to build a robust parser that can simultaneously perform multiple parsing tasks with limited annotated data of eligibility criteria?*” In this work, we aim to investigate the use of a supervised multi-input multi-output (MIMO) sequence labelling model (Jiang et al., 2019) to parse eligibility criteria. This architecture has two modules: a MIMO sequence labelling model, and a self-training method based on heuristic rule correction. In this architecture, multiple input sequences that can be generated automatically include: (1) word embeddings of the original text; (2) part-of-speech tags; (3) language model representation; and (4) concept, attribute, phrase (CAP) tagging. The tag sequences, which must be labelled manually, can be converted into fact and condition tuples jointly (i.e., multiple output). Expressing eligibility criteria in these tuples makes it possible to represent the named entities, temporal constraints (often as conditions), negations, and their relationships in a single universal framework. In this preliminary work, we demonstrate the feasibility of this approach for parsing eligibility criteria with a small labelled dataset.

## 2 Related Work

A number of NLP systems for clinical trial eligibility criteria parsing have been developed previously. These systems can be categorized into (1) rule-based, and (2) machine learning-based systems. Rule-based parsers (e.g., EliXR (Weng et al., 2011), ValX (Hao, Liu, & Weng, 2016), rely on predefined rules, which may not be robust enough to handle complex criteria (e.g., unseen patterns). One the other hand, machine learning-based parsers (e.g., ELiIE (Kang et al., 2017), Criteria2Query (Yuan et al., 2019)) are robust, but require a large training corpus with annotated data to achieve satisfactory performance. Recently, two large manually annotated eligibility criteria datasets were released: the Chia data with 1000 trials (Kury et al., 2020) and the Facebook Research Data with 3314 trials (Tseo, Salkola, Mohamed, Kumar, & Abnousi, 2020). Tian et al. (Tian et al., 2021) recently benchmarked 4 transformers-based NER models on these two datasets and RoBERTa pretrained with MIMIC-III clinical notes and eligibility criteria yielded the highest strict and relaxed F-scores in experiments with both datasets. Further, these existing methods often do not emphasize the representations of the parsing results, leading to difficulty of reusing the annotated training data or the parsing results.

## 3 Methods

### 3.1 Data Source and Data Annotation

#### 3.1.1 Eligibility criteria of Alzheimer’s disease (AD) trials From the *ClinicalTrials*.*gov*, we obtained free-text eligibility criteria of 13 phase III and IV AD clinical trials for existing

Food and Drug Administration (FDA) approved AD drugs.

#### 3.1.2 Annotation process

We followed the tagging schema (i.e., “B/I-XYZ” and “O”) in the original MIMO study (Jiang et al., 2019) to annotate the eligibility criteria, where:

- B: beginning, I: inside;
- X ∈{fact, condition};
- Y ∈{1: subject; 2: relation; 3: object};
- Z ∈{concept, attribute, predicate}.

We decomposed each eligibility criteria into a set of fact and condition tuples. The tags in “B/I-XYZ” format are used for tagging word tokens of each component in the fact and condition tuples, where “B” represents the start word of a tuple component, “I” represents words other than the start word of a tuple component; “X” ∈{f, c}” represent the tuple types of fact (f) and condition (c); both fact and condition tuples are represented by 3 components (1) subject, (2) predicate, and object (3); and “Z” ∈{C, A, P} represent the component roles of concept (C), attribute (A) and predicate phrase (P). Using this format, each word of eligibility criteria can be annotated into 10 different tags as shown in Table 1. Note that any words not in a component of fact and condition tuples are tagged as “O”. An example is shown in Figure 1.

**Table 1:** The examples of “B/I-XYZ” tagging schema.

|  | Fact | Condition |
| --- | --- | --- |
| Subjects | B/I-f1C | B/I-c1C |
| Subject Attributes | B/I-f1A | B/I-c1A |
| Predicates | B/I-f2P | B/I-c2P |
| Objects | B/I-f3C | B/I-c3C |
| Object Attributes | B/I-f3A | B/I-c3A |

**Figure 1.**
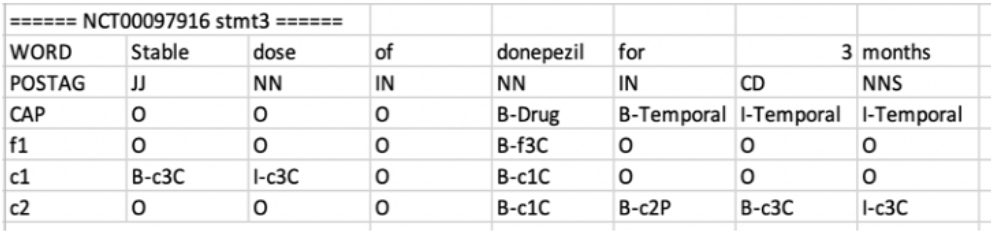
An example of annotating a criterion.

Following this annotation schema, we developed an annotation guideline specially for annotating eligibility criteria. We completed the annotations in multiple rounds, and our annotation process is shown in Figure 2. In each round of annotation, 2 trials were annotated by 2 annotators based on the annotation guideline and Kappa scores were calculated (Glen, 2014). Conflicts between the two annotators were resolved by a third annotator and discussed with the entire study team.

**Figure 2.**
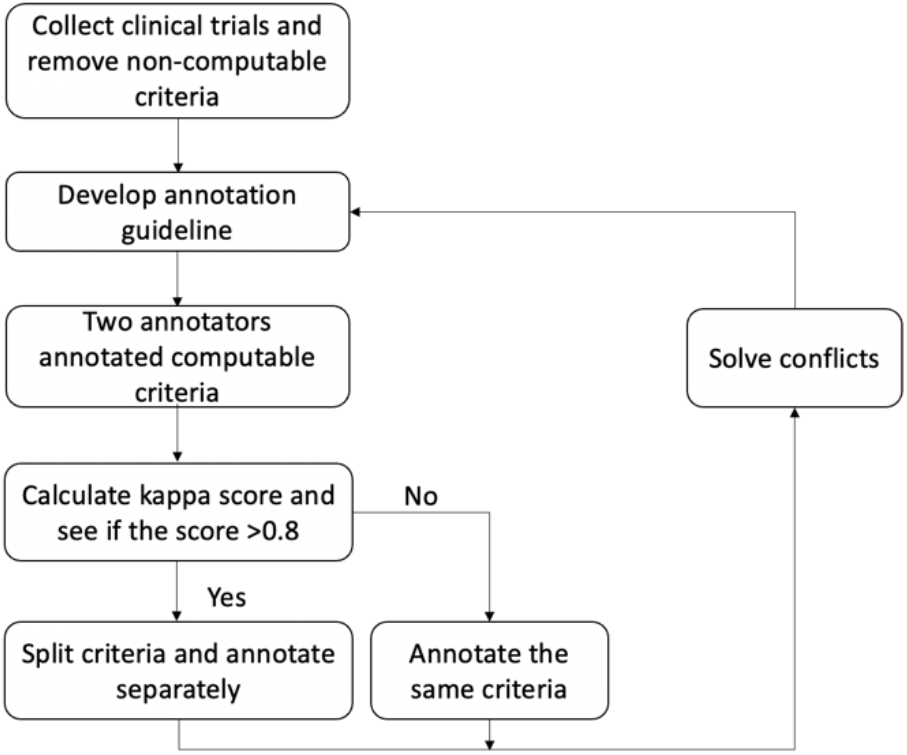
The annotation process.

### 3.2 The Multi-Input Multi-Output Sequence Labeling Model

The multi-input multi-output sequence labeling model, named as MIMO, was proposed by Jiang et al. as a framework for extracting fact and condition tuples from scientific text (Jiang et al., 2019). The advantage of MIMO is that it not only extracts the factual statements (i.e., fact tuples), but also considers the conditions when the fact tuples are true. The MIMO framework has two modules: (1) a multi-input module that takes four input sequences including pre-trained word embeddings, pre-trained language model outputs, part-of-speech (POS) tags, and CAP (i.e., Concepts, Attributes, and Phrases) tags of a sentence and uses a multi-head en-coder-decoder model to generate a sequence representation of the input sentence. The multi-input gates were implemented to control the use of different input sequences (Kaiming He, Zhang, Ren, & Sun, 2016); (2) a multi-output module that takes the sequence representation output of the multi-input module as input and predicts multiple tuple tag sequences for the fact and condition tuples. The multi-output module consists of a tuple component tagging layer, which predicts the tag sequences for fact and condition tuple components, and a tuple completion tagging layer, which predicts multiple tag sequences for the fact tuples and condition tuples. Finally, the complete fact and condition tuples were extracted from the predicted fact and condition tuple tag sequences, respectively, using the matching function as in (Stanovsky, Michael, Zettlemoyer, & Dagan, 2018).

Readers who are interested in the framework can refer to the original paper for more details. The code of the MIMO framework is publicly available at: https://github.com/twjiang/MIMO_CFE.

### 3.3 Evaluation Metrics

We use standard evaluation metrics of precision (P), recall (R) and f1 score (F1) at strict and lenient levels to evaluate performance of the MIMO framework for component tagging and tuple extraction of both fact and condition tuples.

For evaluation of component tagging, each fact or condition tuple component was considered as a named entity. Strict level evaluation requires exact match between the predicted component and the ground truth annotated component for each type of components of fact and condition tuples. Lenient level evaluation requires only overlap between the predicted component and the ground truth annotated component.

For evaluation of tuple extraction, strict level evaluation requires exact match between the extracted tuple and the ground truth annotated tuple for each fact and condition tuple, i.e., each component of the extracted tuple matches exactly each component of the annotated tuple for all 5 different components of subject, subject attribute, predicate, object, and object attribute in each tuple. At the lenient level, an extracted tuple was considered as approximately matched as long as the subject, predicate and object of the extracted tuple overlap with a ground truth annotated subject, predicate and object respectively. Table 2 shows an example illustrating exact and approximate match of tuples:

**Table 2:**
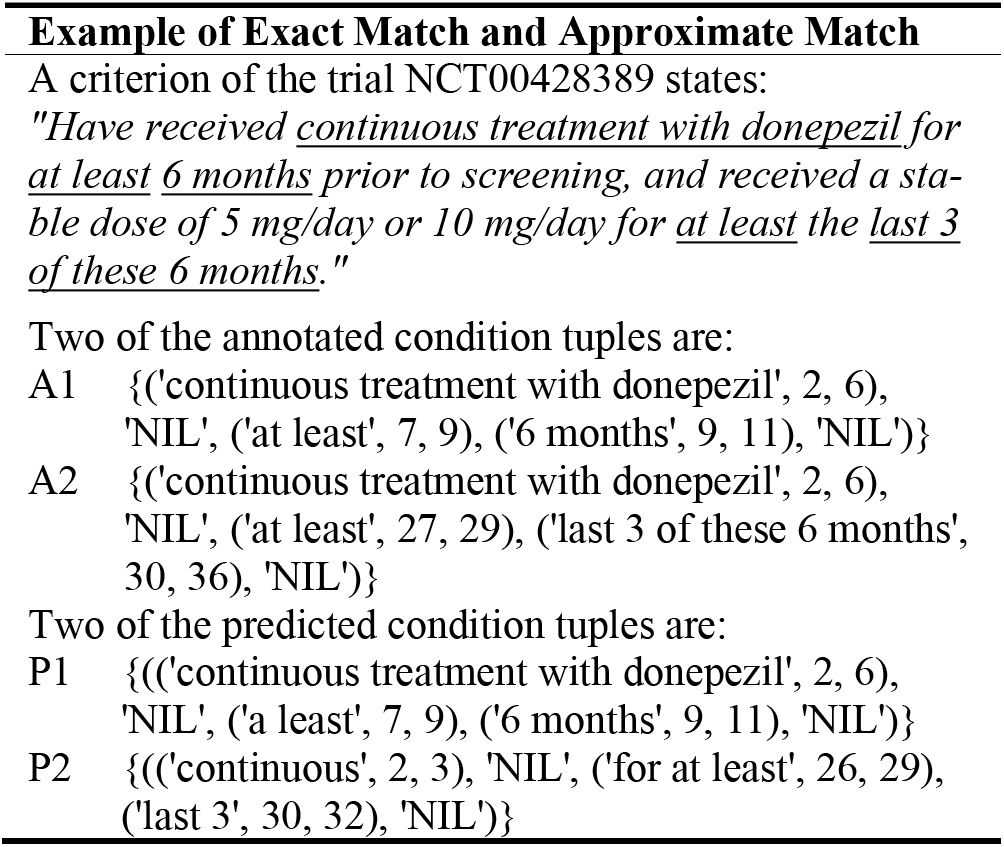
Examples of exact match and approximate match

The predicted tuple P1 is considered as exact match with A1 while P2 is considered as approximate match with A2. Then the precision (P), recall (R) and f1 score (F1) can be calculated using standard formulas based on true positive (TP) (i.e., the number of exact or approximate match tuples), false positive (FP) (i.e., the number of unmatched extracted tuples), and false negative (FN) (i.e., the number of tuples not being extracted).

## 4 Experiment and Results

We conduct the experiments by implementing the MIMO framework with our annotated data and reusing the code made publicly available on GitHub with minor changes to accommodate our workflow and report the results as follows.

### 4.1 Experiment

The MIMO framework include models of different architectures. We selected the MIMO framework with a BERT-based encoder for our experiment because it outperformed frameworks with other architectures as reported by the authors (Jiang et al., 2019).

We split our annotated data by randomly selecting 8 trials as training data and using the remaining 5 trials as test data. The number of tuples and components in the training and test data is given in Table 3. Following the best practice in (Bird, Klein, & Loper, 2009), we used the NLTK (Natural Language Toolkit) package for word tokenization and POS tagging of the input sentences. We obtained the word embeddings with dimension of 50 from the MIMO repository on GitHub. The MIMO framework with a BERT-based encoder uses BERT (Devlin, Chang, Lee, & Toutanova, 2018) as the pre-trained language model. For CAP tagging, we used the NER tags predicted by a NER model based on RoBERTa (Liu et al., 2019), a transformer-based model first pre-trained with general English corpora and further pretrained with MIMIC-III clinical notes (Johnson et al., 2016) and eligibility criteria extracted from more than 350,000 clinical trial summaries on ClinicalTrials.gov. The RoBERTa NER model was then fine-tuned with a dataset derived from Chia, a corpus containing more than 12,000 annotated eligibility criteria from 1,000 Phase IV trials in ClinicalTrials.gov (Tian et al., 2021). We included entities of 6 major types including Condition, Value, Procedure, Drug, Measurement and Temporal in the derived dataset for training the NER model.

**Table 3:**
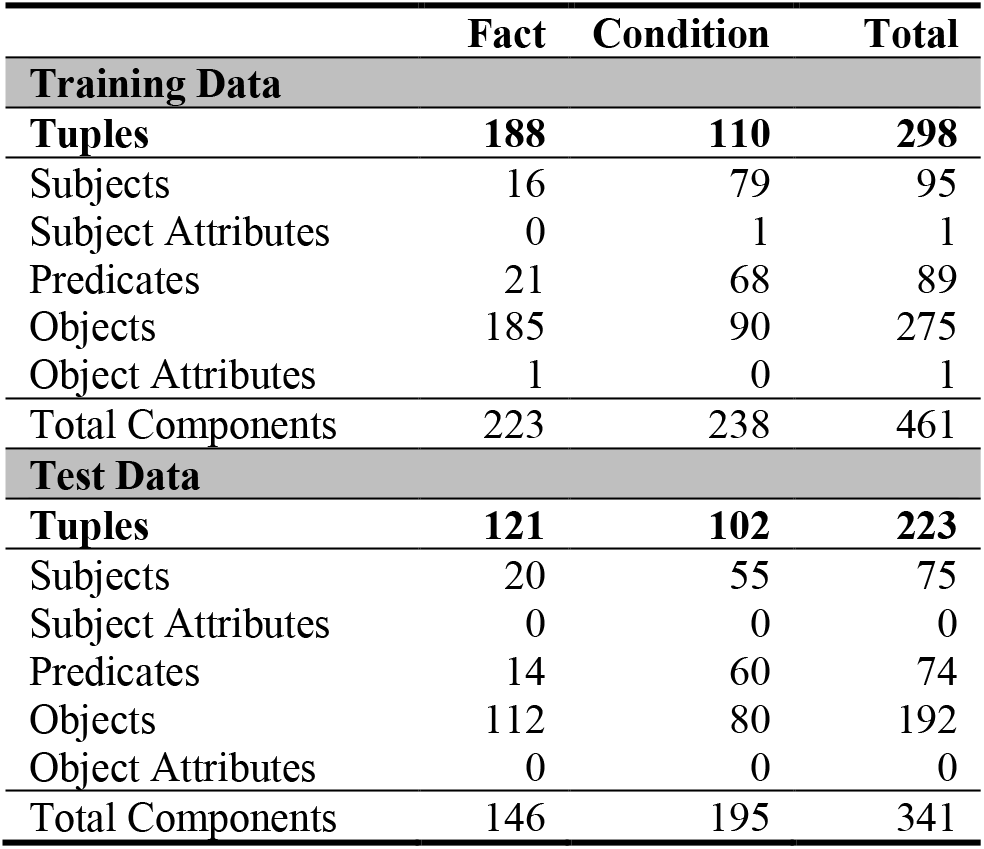
Tuples and Components in Training and Test Data

We used the default hyperparameters set in the MIMO framework and trained the MIMO framework (Jiang et al., 2019) with a BERT-based encoder using different combination of inputs. We experimented with the different sets of input sequences and evaluated the performance using the evaluation metrics described in Section 3.3.

### 4.2 Results

Our experiment results show that the MIMO framework with a BERT-based encoder using all inputs of pre-trained word embeddings, pre-trained language model outputs, POS tags, and CAP tags achieves the best performance in terms of all evaluation measures at strict level for extraction of both fact and condition tuples, and achieves the best performance in terms of precision and f1 score for extraction of fact tuples at the lenient level. Detailed experimental results of the MIMO framework with a BERT-based encoder using all the input sequences are given in Table 4 and Table 5.

**Table 4:**
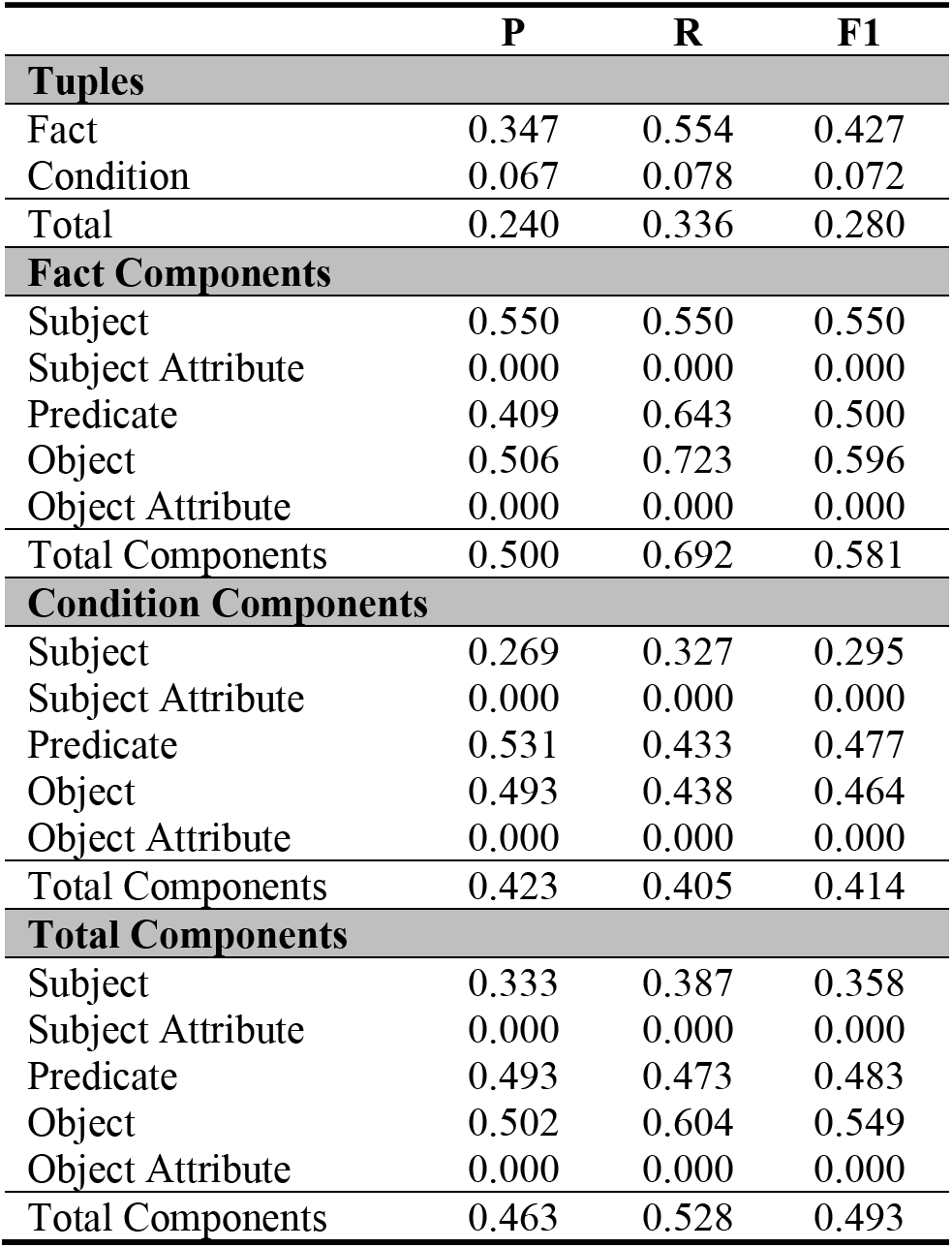
Performance of the MIMO framework with a BERT-based encoder using all inputs at strict level.

|  | P | R | F1 |
| --- | --- | --- | --- |
| **Tuples** |  |  |  |
| Fact | 0.347 | 0.554 | 0.427 |
| Condition | 0.067 | 0.078 | 0.072 |
| Total | 0.240 | 0.336 | 0.280 |
| **Fact Components** |  |  |  |
| Subject | 0.550 | 0.550 | 0.550 |
| Subject Attribute | 0.000 | 0.000 | 0.000 |
| Predicate | 0.409 | 0.643 | 0.500 |
| Object | 0.506 | 0.723 | 0.596 |
| Object Attribute | 0.000 | 0.000 | 0.000 |
| Total Components | 0.500 | 0.692 | 0.581 |
| **Condition Components** |  |  |  |
| Subject | 0.269 | 0.327 | 0.295 |
| Subject Attribute | 0.000 | 0.000 | 0.000 |
| Predicate | 0.531 | 0.433 | 0.477 |
| Object | 0.493 | 0.438 | 0.464 |
| Object Attribute | 0.000 | 0.000 | 0.000 |
| Total Components | 0.423 | 0.405 | 0.414 |
| **Total Components** |  |  |  |
| Subject | 0.333 | 0.387 | 0.358 |
| Subject Attribute | 0.000 | 0.000 | 0.000 |
| Predicate | 0.493 | 0.473 | 0.483 |
| Object | 0.502 | 0.604 | 0.549 |
| Object Attribute | 0.000 | 0.000 | 0.000 |
| Total Components | 0.463 | 0.528 | 0.493 |

**Table 5:** Performance of the MIMO framework with a BERT-based encoder using all inputs at lenient level

|  | P | R | F1 |
| --- | --- | --- | --- |
| **Tuples** |  |  |  |
| Fact | 0.674 | 0.851 | 0.752 |
| Condition | 0.454 | 0.373 | 0.409 |
| Total | 0.590 | 0.632 | 0.610 |
| **Fact Components** |  |  |  |
| Subject | 0.850 | 0.900 | 0.874 |
| Subject Attribute | 0.000 | 0.000 | 0.000 |
| Predicate | 0.500 | 0.714 | 0.588 |
| Object | 0.756 | 0.920 | 0.830 |
| Object Attribute | 0.000 | 0.000 | 0.000 |
| Total Components | 0.738 | 0.897 | 0.810 |
| **Condition Components** |  |  |  |
| Subject | 0.582 | 0.582 | 0.582 |
| Subject Attribute | 0.000 | 0.000 | 0.000 |
| Predicate | 0.776 | 0.600 | 0.677 |
| Object | 0.747 | 0.563 | 0.642 |
| Object Attribute | 0.000 | 0.000 | 0.000 |
| Total Components | 0.695 | 0.580 | 0.632 |
| **Total Components** |  |  |  |
| Subject | 0.644 | 0.667 | 0.655 |
| Subject Attribute | 0.000 | 0.000 | 0.000 |
| Predicate | 0.690 | 0.622 | 0.654 |
| Object | 0.753 | 0.771 | 0.762 |
| Object Attribute | 0.000 | 0.000 | 0.000 |
| Total Components | 0.717 | 0.716 | 0.716 |

From our experiment, we observed that the MIMO framework tended to extract more tuples than the annotated goldstandard. This brings higher recall but lower precision in most of the experiments. Another observation is that the MIMO framework achieved better performance for fact tuple extraction than performance for condition tuple extraction. One of the reasons for this may be because condition tuples in eligibility criteria of clinical trial summaries are more complicated than fact tuples. In addition, the small sample size of the annotated data from only 13 trial summaries may not be adequate for training a deep learning model to achieve a good performance.

## 5 Discussion and Conclusions

In this preliminary work, we evaluated the feasibility of using the MIMO framework to parse clinical trial eligibility criteria. Using 13 AD trials, we achieved a reasonable performance in terms of lenient-level F1 for recognizing components of fact (0.81) and condition tuples (0.72), respectively and then the entire tuples (0.61). The reason for the lower performance of condition tuples could be attributed to the small sample size. And the unsatisfactory performance of the strict-level evaluation is mainly due to inaccurate tuple components extraction. Nevertheless, representing eligibility criteria into logical and semantically clear fact and condition tuples can potentially make subsequent translation of these tuples into database queries more reliable. In future work, we will refine the annotation guideline and annotate more trials to increase the training samples. We will also integrate the results with the entity type recognition model (RoBERTa-MIMIC-Trial) that we previously built (Tian et al., 2021), which can potentially improve the model performance. We will explore ways of building database queries against realworld EHR data using the tuples and evaluate cohort identification performance.

## Data Availability

All data produced in the present study are available upon reasonable request to the authors.

## Acknowledgments

This study was supported in part by the National Institutes of Health (NIH) under awards R21AG061431, R21AG068717, R21 CA253394, and UL1TR001427.

